# Uncharacterized *MYH9* Germline Mutations in a Microcystic Adnexal Carcinoma Mimicker: Benign Deep Syringoid Ductal Proliferation (BDSDP) with Elastic Fiber Aggregation

**DOI:** 10.1101/2025.08.03.25332336

**Authors:** Etta Hanlon, Katherine L. Brown, Haley M. Michel, Clinton Roby, Matthew G. Urbano, Dzenis Mahmutovic, Priyenka Khatiwada, Jane Gay, Anne M. Brown, Douglas J. Grider, Carla V. Finkielstein

**Author notes:** Corresponding Authors: Douglas J. Grider, MD, Carla V. Finkielstein.

## Abstract

**Importance:** Microcystic adnexal carcinoma (MAC) is a rare, locally aggressive sweat gland neoplasm sometimes misdiagnosed due to its histologic similarities to benign adnexal proliferations. MYH9-associated elastin aggregation syndrome (MALTA) is an inherited condition characterized by benign MAC-like ductal lesions and by abnormal elastic fiber deposition.

**Objective:** To report previously uncharacterized heterozygous germline mutations in the *MYH9* gene in a patient presenting benign deep syringoid ductal proliferations and papillary dermal elastic fiber aggregation.

**Design, Setting, Participants:** Clinical report with genetic and structural analysis. Dermatology outpatient. A male in their 20’s presenting with long-standing, stable erythematous nodules on the right infraorbital region and left zygomatic arch. Genetic testing of first-degree relatives and structural simulations were performed to assess variant impact.

**Main Outcomes and Measures:** Histological evaluation of the patient’s lesions revealed benign deep syringoid ductal proliferations with papillary dermal elastic fiber aggregation, distinguishing them from microcystic adnexal carcinoma. Germline genetic testing identified three heterozygous *MYH9* variants, two previously uncharacterized, all showing Mendelian segregation in first-degree relatives and associated with structural rearrangement.

**Results:** Histologic evaluation of the facial lesions revealed keratin-filled microcysts and deep dermal and subcutaneous cords with ductal structures resembling MAC. Immunohistochemistry showed apocrine differentiation (EMA+/CD15+/GCDP+) and basaloid myoepithelial cells positive for p63. No evidence of perineural invasion was observed. Elastic tissue staining showed dense, ball-like aggregates of elastic fibers in the papillary dermis. Germline testing identified c.1363G>A (p.Gly455Ser) in the myosin head domain, and c.4490G>A (p.Arg1497Gln) and c.4876A>G (p.Ile1626Val) in the tail domain of Myosin-9. Saliva-based testing confirmed Mendelian segregation in multiple first-degree relatives. Missense mutations were predicted to alter the coiled-coil structure, potentially disrupting chain interactions and affecting the motif’s parallel versus antiparallel orientation.

**Conclusions and Relevance:** This case broadens the phenotypic and genotypic spectrum of MALTA syndrome and introduces the diagnostic term: benign deep syringoid ductal proliferation (BDSDP) with elastic fiber aggregation. The findings underscore the diagnostic challenges in distinguishing BDSDP from MAC and highlight the critical role of integrating histopathologic, immunohistochemical, and genetic data in accurate diagnosis. These results support the need for further investigation into MYH9-associated adnexal neoplasia and its underlying molecular mechanisms.

**Key Points:** *Question:* How do germline *MYH9* variants contribute to the pathogenesis of benign deep syringoid ductal proliferations with elastic fiber aggregation, a phenotype that clinically and histologically mimics microcystic adnexal carcinoma?

*Findings:* Genetic analysis revealed two previously unreported heterozygous variants in the *MYH9* gene: c.1363G>A (p.Gly455Ser), located in the myosin head domain, a region previously associated with MALTA syndrome, and a variant in the myosin tail domain, c.4490G>A (p.Arg1497Gln). A third mutation, c.4876A>G (p.Ile1626Val), was also detected. All three variants demonstrated Mendelian segregation from the parents, were identified in multiple family members, and were predicted to cause structural perturbations.

*Meaning:* These findings provide strong evidence for a heritable contribution of these mutations to the observed phenotype. The presence of these *MYH9* variants highlights a potential functional impact on protein structure and activity. This pattern supports the hypothesis that *MYH9* mutations may underlie or modify the pathogenesis of benign syringoid ductal proliferations, expanding the known spectrum of MYH9-associated conditions and offering a molecular basis for improved diagnosis and familial risk assessment.

## Introduction

Microcystic adnexal carcinoma (MAC) is a rare, locally aggressive sweat gland tumor that can be misdiagnosed due to histologic overlap with benign adnexal lesions ^1^. It typically presents as a firm head or neck nodule with deep infiltration, keratin cysts, and perineural invasion ^2^. Benign “MAC mimickers,” such as syringomas and inherited syndromes, resemble MAC histologically but lack invasive features ^2–4^. In 2019, Fewings et al. unified two inherited conditions as MALTA (*MYH9*-associated elastin aggregation syndrome), an autosomal dominant disorder caused by germline *MYH9* mutations and characterized by elastic fiber aggregation and benign ductal proliferations ^4,5,6^. Unlike other MYH9-related conditions such as MATINS or DFNA17, MALTA manifests solely with cutaneous findings ^5,7^. Accurate diagnosis is critical to prevent overtreatment.

Evidence implicates *MYH9* in skin carcinogenesis, expanding its clinical relevance beyond hematologic and auditory syndromes. Accordingly, a *MYH9* mutation in the myosin head domain has also been linked to MALTA syndrome and early-onset squamous and basal cell carcinomas ^5^. We report a case of benign ductal proliferation with elastic fiber aggregates and MYH9 mutations segregating in a Mendelian pattern. Structural modeling suggests these variants disrupt Myosin-9 assembly and stability, supporting MALTA as a distinct cutaneous syndrome with potential oncogenic relevance.

## Methods

Biopsy samples from the patient’s facial lesions and saliva from the patient and their direct family members were collected for genomic and histopathological analysis. A benign deep syringoid ductal proliferation (BDSDP) specimen served as the primary genomic sample, with an angiofibroma used as control. Forty coding exons of the *MYH9* gene were amplified and sequenced (^8^ and **eTable 1**). Variants were analyzed using Sequencher and assessed for pathogenicity through VarSome (**eMethods**).

Myosin-9 structure was modeled using AlphaFold and visualized in PyMOL (**eMethods**). AlphaMissense scores were used to evaluate the pathogenic potential of identified missense mutations. Further, DynaMut2 in silico modeling was used to assess the impact of mutations on Myosin-9 stability and conformational dynamics (**eMethods**). Myosin-9 wild-type and mutant dimers were modeled using AlphaMultimer and validated (**eMethods**).

## Results

A male in their 20’s presented with two longstanding facial nodules initially concerning for microcystic adnexal carcinoma (MAC) (**Figure 1**A). Histologic evaluation revealed ductal structures lacking perineural invasion, with p63+ myoepithelial cells and elastic fiber aggregation in the papillary dermis, supporting a newly defined diagnosis of benign deep syringoid ductal proliferation (BDSDP). Immunohistochemistry showed focal GCDFP-15+/EMA+ expression along the inner ductal surfaces, while CD15+ indicated apocrine rather than eccrine differentiation (**Figure 1**B-C). Together, the EMA+/CD15+/ GCDFP-15+ profiles supported the revised diagnostic classification.

**Figure 1.**
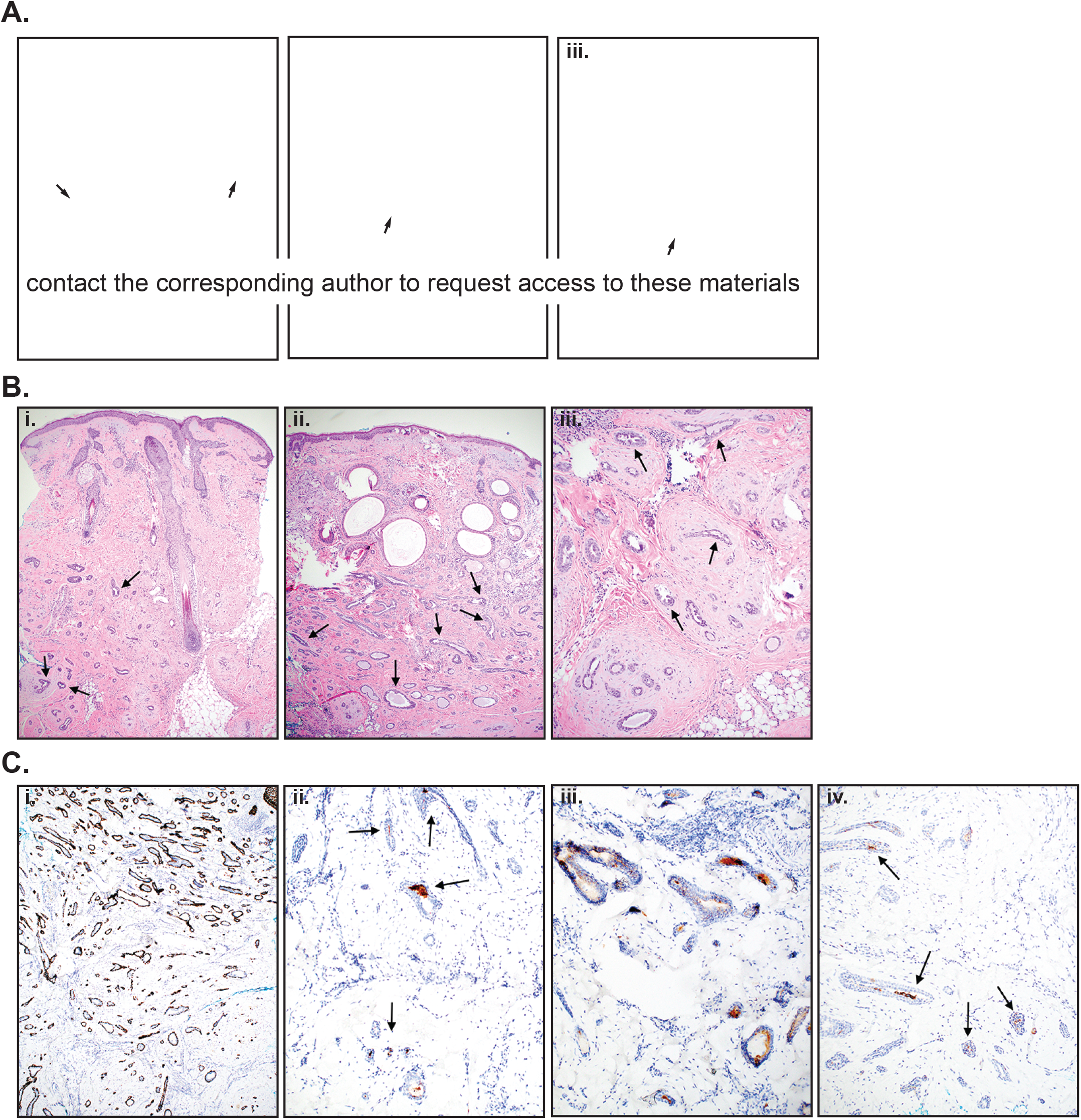
Clinical presentation and histopathological features. (**A**) Clinical photograph (i) showing nodular erythematous swellings on the right infraorbital region (ii) and left zygomatic arch (iii), measuring approximately 2.5 cm and 3.2 cm in diameter, respectively. (**B**) Hematoxylin and eosin (H&E) stained tissue sections front the right infraorbital region at 4x (i), with arrows indicating ducts exhibiting apocrine-like changes. H&E stained tissue sections from the left zygomatic arch lesion at 4x (ii) and 10x (iii) reveal keratin horn cysts in the upper dermis, a feature mimicking microcystic adnexal carcinoma (MAC), along with myoid/collagenous stroma surrounding the cords and ductular proliferation, which are not typical of MAC. Arrows mark ducts with features of apocrine differentiation. (**C**) Immunohistochemical ancillary studies support apocrine differentiation (i) p63 staining highlights the nuclei of the basal myoepithelial cell layer of the infiltrative ducts (10x), (ii) GCDFP-15 focally marks the luminal duct surface, a feature sometimes seen with apocrine differentiation (10X), (iii) EMA marks the luminal surface of the infiltrative glandular duct structures (10X), (iv) CD15 positivity along the lumen surface and within the infiltrative ducts favors apocrine over eccrine origin (10X).

Given the unusual bilateral presentation and young age, germline testing was pursued. Sequencing of the *MYH9* gene identified heterozygous missense mutations: c.1363G>A (p.Gly455Ser) and c.4490G>A (p.Arg1497Gln) as well as c.4876A>G (p.Ile1626Val), all confirmed in both lesions and saliva samples, consistent with a germline origin (**Figure** 2A-B). Eight intronic variants, including two uncharacterized SNPs; were identified but none were predicted to affect splicing or gene function (**eTable 2**). The Gly455Ser variant in Myosin-9 was predicted to be “pathogenic strong” by *in silico* analysis ^9^. Family testing showed paternal inheritance of c.1363G>A (p.Gly455Ser) and c.4876A>G (p.Ile1626Val) (in cis) and maternal inheritance of c.4490G>A (p.Arg1497Gln), resulting in compound heterozygosity in both affected siblings (**Figure 2**B-D). Unlike previously reported MALTA and MATINS syndromes (**Figure 2**E), this pedigree uniquely suggests an autosomal recessive inheritance pattern and supports the hypothesis that structurally disruptive mutations in the Myosin-9 motor domain contribute to the pathogenesis of MALTA syndrome.

**Figure 2.**
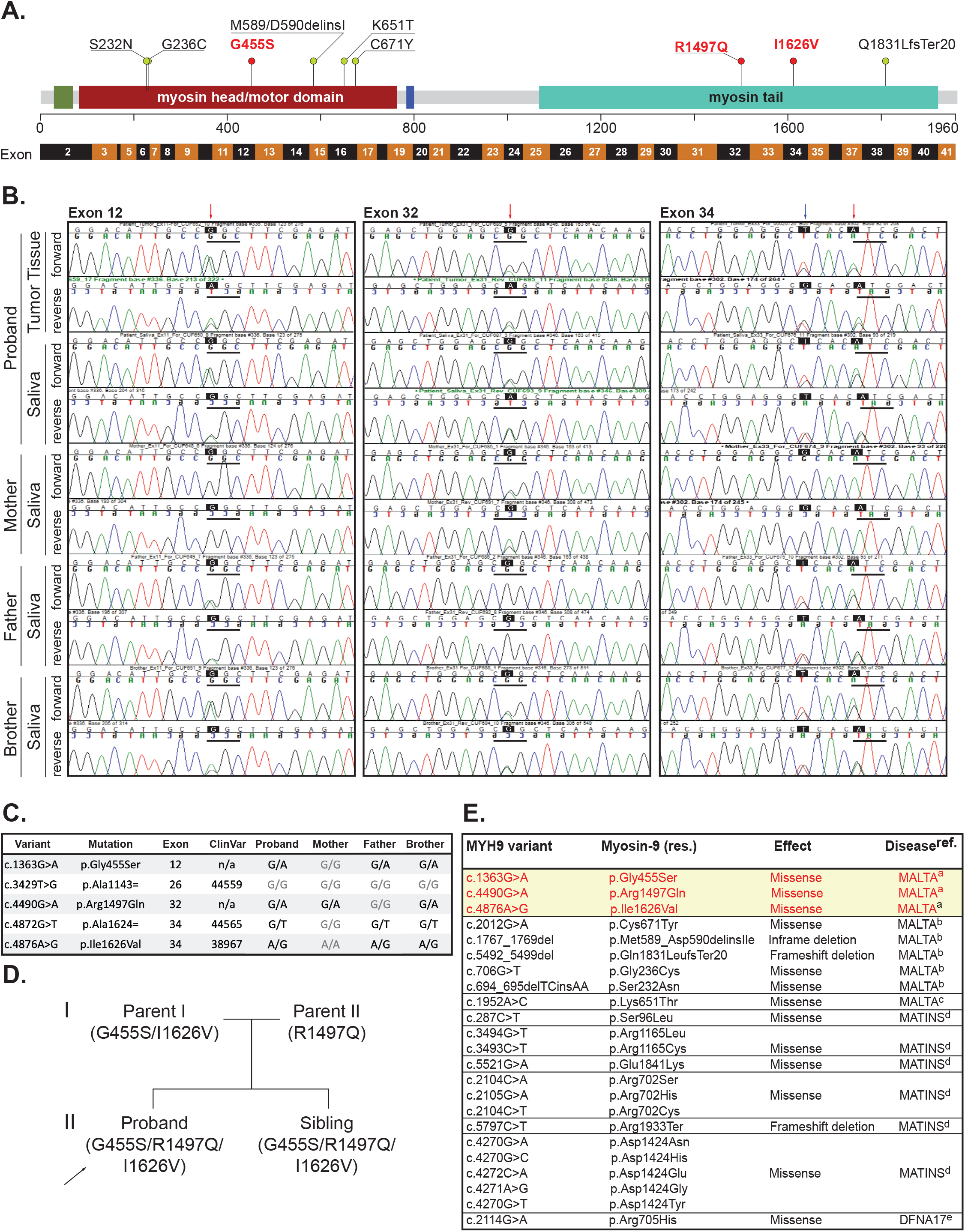
Genomic identification of familial *MYH9* variants. (**A**) Scaled schematic representation of the full-length Myosin-9 (residues 1–1960), showing structural and functional domains: myosin N-terminal SH3-like domain (green), myosin head/motor domain (maroon), IQ calmodulin-binding motif (blue), and myosin tail (cyan). Previously reported pathogenic mutations (missense, deletions/insertions, and frameshifts) linked to MALTA are shown in green lollipops; red lollipops indicate newly identified variants. Below, a schematic of the 41 *MYH9* exons, of which 40 are coding, is aligned with the corresponding protein regions. (**B**) Representative sequencing chromatograms of gDNA from tumor and saliva samples from the proband and immediate family members. Detected variants are shown: c.1363G>A (p.Gly455Ser; exon 12), c.4490G>A (pArg1497Gln; exon 32), and c.4876A>G (pIle1626Val; exon 34). Black boxes highlight the variant positions; red arrows denote altered peak in the chromatogram; blue arrow indicates a synonymous variant in exon 34 (c.4872G>T; p.Ala1624=). Codon positions are marked with solid black lines. Double peaks indicate heterozygosity. The *MYH9* c.1363G>A (p.Gly455Ser), c.4490G>A (p.Arg1497Gln), and c.4876A>G (p.Ile1626Val) mutations were confirmed as heterozygous in both normal tissue and saliva indicating these are constitutional mutations rather than a somatic change confined to the tumor. (**C**) Table summarizing the *MYH9* variants identified in gDNA from saliva samples of all individuals in the pedigree, including corresponding protein changes, exon locations, and ClinVar accession numbers (if available). For each variant, the genotype (e.g., G/A, G/G, G/A, G/T, A/G) is provided for the Proband and first-degree relatives. (**D**) Pedigree chart illustrating inheritance patterns of three *MYH9* variants across two generations (I and II). Genomic DNA from saliva samples revealed that the parent (I-1) carries the Gly455Ser and Ile1626Val mutations on the same allele, while the other parent (I-2) carries the Arg1497Gln variant. The chart shows a Mendelian segregation pattern, with parent inheritance of Gly455Ser and Ile1626Val and Arg1497Gln in both offspring (II-1 and II-2). The proband (II-1) is indicated by an arrow. Variant distribution is shown using patterned fills: solid black for Gly455Ser and Ile1696Val, and diagonal stripes for Arg1497Gln. (**E**) Summary of *MYH9* variants associated with MALTA (MYH9-associated elastin Aggregation), MATINS (Macrothrombocytopenia and granulocyte inclusions with or without nephritis or sensorineural hearing loss), and DFNA (Deafness, autosomal dominant 17) phenotypes, including those located in the coding region and reported in this work. References (a) this work, (b) ^4^, (c) ^5^, (d) only included 6 hotspot mutations of 107 MATINS mutations reported ^15^, (e) ^7^.

To test this hypothesis, we mapped all motor domain mutations onto the AlphaFold-predicted Myosin-9 structure, revealing that they cluster within a topologically confined region of the protein, suggesting structural or functional convergence (**Figure 3**A, upper and middle panels) ^10,11^. AlphaMissense ^12^ high pathogenicity scores indicated that these mutations, particularly Gly455Ser in the motor domain, likely destabilize Myosin-9 structure (**Figure 3**A, lower panels). All variants demonstrated negative ΔΔG values consistent with destabilizing effects on Myosin-9 (**Figure 3**B, top table) ^13^. Gly455Ser, in particular, caused local structural rearrangements, introducing a new hydrogen bond with Ser176 and disrupting interactions with nearby residues (**Figure 3**B, bottom panels). These changes alter local packing, supporting a direct mechanism for pathogenicity by effectively reshaping the interaction landscape within the motor domain (**Figure 3**B, bottom panels).

**Figure 3.**
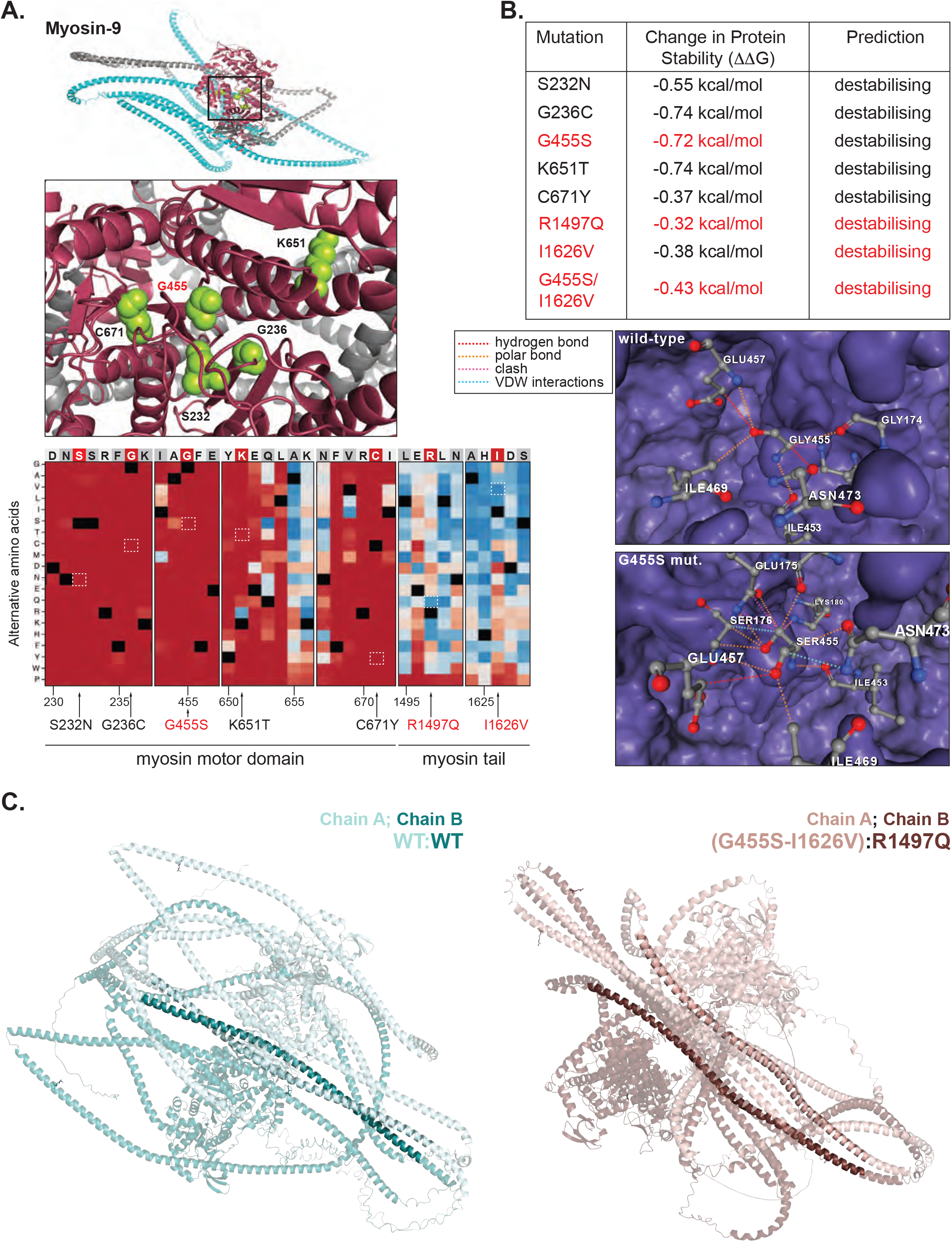
*In silico* characterization of MHY9 variants. (**A**) Cartoon representation of AlphaFold-predicted Myosin-9 structure, with domains color-coded as in (Figure 2A). The boxed region within the head/motor domain is shown enlarged in the middle panel. Residues previously mutated in MALTA patients are shown as green spheres. The bottom panels display the AlphaMissense pathogenicity heatmap for the amino acid region surrounding the wild-type residue (highlighted by a red box in the top sequence). The corresponding mutated substitution is indicated by a dashed white box. Unlike mutations in the tail domain that received low pathogenicity scores (Arg1497Gln: 0.126 and Ile1626Val: 0.086), residues Ser232Asn, Gly236Cys, Gly455Ser, Lys651Thr, Arg1497Gln, and Cys671Tyr showed high pathogenicity scores (0.999, 0.998, 0.997, 0.983, 1.000, respectively), supporting the interpretation that these variants likely disrupt MYH9 function. The heatmap color gradient represents AlphaMissense pathogenicity scores, ranging from blue (low probability of pathogenicity, score near 0) to red (high probability of pathogenicity, score near 1). **(B)** DynaMut2 predictions of the thermodynamic impact of various *MYH9* mutations. The table shows ΔΔG values, with mutations identified in this study highlighted in red. The bottom panels display surface and stick models of residue interactions (hydrogen bonds, polar, and Van der Waals forces) surrounding wild-type Gly455 (top) and mutant Ser455 (bottom), revealing local structural disruptions. Notably, Ser455, but not Gly455, can form a hydrogen bond with Ser176. Additional changes in nearby interactions, including those involving Asn473 and Ile453, suggest a broader effect on the local structural environment due to the Gly455Ser substitution. (**C**) AlphaFold-Multimer de novo models of the wild-type MYH9 homodimer (left) and the patient-derived mutant dimer (right) are shown. In the wild-type model, chain A (darkt teal) and chain B (light teal) include residues Gly455, Arg1497, and Ile1626. In the mutant model, residues were assigned based on familial segregation: chain A (light brown) contains mutations Ser455 and Val1626, while chain B (dark brown) includes the Gln1497 variant.

AlphaMultimer models were generated for wild-type (WT:WT) and mutant [(G455S-I1626V):R1497Q] Myosin-9 dimers (**eFigures 1–2**). Both showed similar myosin head positioning, but WT:WT retained a more complete coiled-coil motif, whereas the mutant lacked coiled-coil interactions near I1626V (**Figure 3**C). *In silico* mutagenesis predicted reduced homodimer stability for all variants (positive ΔΔG), with the (G455S-I1626V):R1497Q combination showing the greatest destabilization (**eFigure 2**B). I1626V disrupted hydrophobic packing and coiled-coil integrity, consistent with prior findings on Ile→Val substitutions reducing coiled-coil stability ^14^. Together, these findings suggest that *MYH9* mutations contribute to MALTA syndrome by disrupting polymerization and cytoskeletal regulation, potentially driving elastic fiber aggregation, and suggest an autosomal recessive inheritance pattern.

## Discussion

This study expands the clinical and molecular landscape of MALTA syndrome by identifying novel *MYH9* mutations with predicted structural effects. Germline testing, segregation analysis, and in silico modeling support their pathogenicity and suggest a previously unrecognized autosomal recessive inheritance. These findings underscore the diagnostic challenges of MAC mimickers and the importance of multidisciplinary approaches in characterizing inherited skin disorders.

*MYH9* mutations cause a range of disorders by affecting either the motor domain (ATP hydrolysis, actin binding) or the tail domain (filament assembly, intracellular transport). For example, Arg702Cys disrupts electrostatic interactions critical for ATPase activity, while Asp1424Asn impairs filament assembly at the dimer interface, both linked to severe MATINS phenotypes with systemic features like macrothrombocytopenia, nephropathy, or deafness□^15^. In contrast, *MYH9* variants in MALTA syndrome primarily affect the skin. Structural integrity of coiled-coil motifs relies on hydrophobic residues and proper side chain packing. In our study, Gly455Ser introduces polarity into the motor domain, disrupting local packing, and Ile1626Val alters the tail domain’s hydrophobic core, potentially destabilizing the dimer. Similar coiled-coil–disrupting Myosin-9 tail mutations have been reported in MALTA and related disorders□^4^.

Genetic analysis revealed that the affected siblings inherited Gly455Ser and Ile1626Val in cis from one parent and Arg1497Gln from the other parent, resulting in compound heterozygosity. This inheritance pattern contrasts with previously reported dominant cases of MALTA and MATINS and supports a recessive model. Structural modeling further supports this interpretation, showing that both Gly455Ser and Ile1626Val cause domain-specific destabilization that may act synergistically to drive the phenotype only when co-inherited. The absence of symptoms in heterozygous family members reinforces this model. These findings support the need for continued dermatologic monitoring and underscore the importance of integrating molecular and histopathologic data when evaluating adnexal tumors and assessing the impact of MYH9 mutations on skin pathology.

### Limitations

This study offers consistent clinical, genetic, and structural evidence linking Myosin-9 mutations to MALTA syndrome, though some limitations remain. As a single-family case report, the generalizability of the findings is limited, and the incomplete penetrance observed in an asymptomatic sibling suggests additional genetic or environmental modifiers. While in silico models predict disruptive structural consequences for the identified variants, these effects were not validated through functional assays. Furthermore, the absence of longitudinal clinical data and tissue-level protein analyses limits insight into disease progression and molecular pathology.

## Conclusions

By identifying two previously uncharacterized *MYH9* mutations, this work links specific structural perturbations in Myosin-9 to a benign dermatologic presentation with potential oncogenic implications. Integrated dermatologic, histologic, genetic, and structural data reveal a mechanistic basis for pathogenicity and suggest a previously unrecognized autosomal recessive inheritance in a condition typically considered dominant. These findings underscore the value of combining molecular and histologic data to diagnose MAC mimickers and inherited adnexal tumors.

## Data Availability

All data produced in the present study are available upon reasonable request to the authors

## Author Contributions

Hanlon had full access to all of the data in the study and takes responsibility for the integrity of the data and the accuracy of the data analysis. Concept and design: Hanlon, K. Brown, Grider, Finkielstein. Acquisition, analysis, or interpretation of data: Hanlon, K. Brown, Michel, A. Brown, Roby, Urbano, Mahmutovic, Khatiwada, Gay, Grider, Finkielstein. Drafting of the manuscript: Hanlon, Grider, Finkielstein. Critical review of the manuscript for important intellectual content: Hanlon, K. Brown, A. Brown, Roby, Urbano, Mahmutovic, Grider, Finkielstein. Statistical analysis: Finkielstein, A. Brown. Obtained funding: Finkielstein. Administrative, technical, or material support: Hanlon, Roby, Urbano, Mahmutovic. Supervision: Grider, Finkielstein.

