## Supplementary Material for "Uncharacterized *MYH9* Germline Mutations in a Microcystic Adnexal Carcinoma Mimicker: Benign Deep Syringoid Ductal Proliferation (BDSDP) with Elastic Fiber Aggregation"

**eMethods**

**Tissue collection and histopathological evaluation.** Biopsy samples were collected from the left temple, right cheek, and right nostril following consultation with a board-certified dermatopathologist (DJG). Tissues were fixed in 10% neutral buffered formalin, paraffin-embedded, and processed for genomic and ancillary immunohistochemical analyses. Initial histopathologic evaluation was performed on 3 μm sections stained with hematoxylin and eosin. Immunohistochemistry was performed on formalin-fixed, paraffin-embedded tissue sections using antibodies against GCDFP-15, EMA, and CD15 to assess for apocrine differentiation. Staining patterns were evaluated for characteristic features such as apical decapitation secretion, best seen on the H&E stained tissue sections, and membrane positive staining for GCDFP-15, EMA and CD15, in ductal structures. In addition, p63 immunohistochemistry stained the nuclei of basal myoepithelial cells lining the outer wall of the ductal structures. The left temple biopsy material was used for genomic analysis, while an angiofibroma from the right nostril served as a control. For DNA extraction, 10 μm sections were cut from each paraffin block. Saliva samples were collected from the patient and relatives using Disposable Sampler Saliva Collection Kits (Wuxi NEST Biotechnology Co., Wuxi, Jiangsu, China). The patient provided informed consent for the use of samples in research studies and publication.

**Genomic DNA purification, amplification, and sequencing.** Genomic DNA (gDNA) was extracted from FFPE samples from the zygomatic arch lesion and from saliva to confirm the presence of germline mutations, using QIAamp DNA FFPE Advanced kit (QIAGEN, Hilden, Germany) and the Monarch® Spin gDNA Extraction kit (New England Biolabs, Ipswich, MA), respectively. Amplification of all 41 exons from *MYH9* was performed using gDNA as template and the *Power* SYBR^TM^ Green kit (Applied Biosystems, Thermo Fisher Scientific, Waltham, MA), with the following cycling program: 95ºC 10 min; 45 cycles of 95ºC for 15 sec, 58ºC for 20 sec, and 60ºC for 75 sec; followed by 65ºC for 2 min and a melt curve from 65 to 95ºC. Primers used for amplification included those reported by Kunishima et al ^8^, along with additional primers (eTable 1) designed using the PrimerQuest™ Tool (Integrated DNA Technologies, Inc., San Diego, CA). Amplicons were treated with ExoSAP-IT™ PCR Product Cleanup Reagent (Applied Biosystems, Foster City, CA) according to the manufacturer’s protocol and submitted to Eurofins Genomics (Louisville, KY) for Sanger sequencing. Resulting sequence data were analyzed using Sequencher v5.4.6 software (Gene Codes Corporation, Ann Arbor, MI). Identified variants were evaluated for frequency and clinical relevance using VarSome (Saphetor, Lausanne, Switzerland)^9^.

***MYH9* gene structure and model prediction.** The human *MYH9* gene (Chr. 22) transcribes a 7,451 bp full-length mRNA (NM_002473), comprising 41 exons, 40 of which are coding, and encodes a canonical protein of 1,960 amino acids known as non-muscle myosin heavy chain II A (NMHC IIA), also referred to as Myosin-9 (UniProt P35579). The domain architecture of Myosin-9 includes an ATP-binding motif (residues 174 to 181), a myosin N-terminal SH3-like domain (residues 27 to 77), a myosin motor domain (residues 83 to 764), an IQ (Ile/Gln) domain (residues 783-799), typically found in proteins regulated by calcium/calmodulin signaling, and a myosin tail domain (residues 1067 to 1923).

The Myosin-9 structure was predicted using AlphaFold3 ^10^ and retrieved via UniProt (accession# P35579). The resulting model (AF-P35579-F1) showed high confidence scores across most of the sequence, with per-residue predicted Local Distance Difference Test (pLDDT) values above 90, indicating a highly reliable structural prediction. Accordingly, the highest-confidence regions were primarily located in protein-protein binding domains and well-structured coiled-coil regions, consistent with high-resolution crystallographic data (PDB identifiers 3ZWH, 4CFQ, 4CFR, 4ETO). Three short, disordered regions, residues 1035-1057, 1118-1137, and 1877-1960, were identified based on lower pLDDT scores. All structural visualizations and graphs were generated using PyMOL (The PyMOL Molecular Graphics System, Version 2.5, Schrödinger, LLC).

**AlphaMissense pathogenicity prediction for *MYH9* variants in Myosin-9.** Missense mutations in Myosin-9, including Ser232Aspn, Gly236Cys, Gly455Ser, Lys651Thr, Arg1497Gln, and Cys671Tyr, were evaluated using the AlphaMissense Pathogenicity Heatmap, which integrates AlphaFold structural data with evolutionary and functional features ^12^ Each substitution is scored from 0 (pathogenic benign; blue in the heatmap) to 1 (highly pathogenic; red in the heatmap). These specific mutations received high pathogenicity scores, suggesting they affect structurally or functionally important regions of the protein.

***In-silico* analysis of protein stability.** DynaMut2 was used to assess the structural and thermodynamic effects of missense mutations in Myosin-9 ^13^. This tool combines normal mode analysis and graph-based signatures to evaluate how single-point mutations influence protein stability and conformational flexibility. For each variant, DynaMut2 predicts the change in Gibbs free energy (ΔΔG, in kcal/mol) upon mutation. Mutations yielding ΔΔG values < 0 were interpreted as destabilizing (i.e., leading to reduced protein stability), while those with ΔΔG > 0 were classified as stabilizing. Structural analysis was performed using the AlphaFold-predicted Myosin-9 (UniProt P35579) as input. Outputs included both ΔΔG values and graphical representations of altered residue interactions, including hydrogen bonds, Van der Waals forces, and electrostatic contacts, comparing wild-type and mutant structures.

**Modeling of Myosin-9 dimers.** Homodimeric models of human wild-type (UniProt: P35579, <https://www.uniprot.org/uniprotkb/P35579/entry>) and mutant Myosin-9 were generated using AlphaFold-Multimer (v2.3.2) ^1,2^. Amber minimization was applied to the wild-type model, but not to the (Gly455S-Ile1626V):Arg1497Gln [also (G455S-I1626V):R1497Q] mutant due to computational resource constraints. Structural evaluation was performed using SWISS-Model was used to evaluate stereochemistry and to estimate global and local model quality of the top models ranked by AlphaFold2 ^3^. These models were further analyzed using the Schrödinger Suite (Schrödinger Release 2025-2: Prime, Schrödinger, LLC, New York, NY, 2025). Residue scanning was performed on the wild-type Myosin-9 homodimer to determine changes in free energy (ΔΔG) associated with the G455S, I1626V, and R1497Q mutations, focusing on their impact on homodimer stability and interchain binding affinity. The mutations were modeled simultaneously, with G455S and I1626V occurring on Chain A and R1497Q on Chain B. Changes in structural stability (ΔΔGstability) were calculated as the difference in free energy between the unfolded and folded states of the wild-type and mutant dimers. Changes in binding affinity (ΔΔGbind) between dimer chains were computed as the difference in free energy change between the wild-type and mutant homodimer monomers. Both ΔΔGstability and ΔΔGbind were estimated using Schrödinger Prime MM-GBSA ^4,5^. Changes in hydropathy for each mutation were reported based on the Kyte–Doolittle scale to indicate shifts in hydrophobicity or hydrophilicity ^6^. Model quality metrics indicate moderate confidence, with Ramachandran plots supporting the predicted secondary structure (**eFigure 2**, panels C and I). Although deviations from ideal geometry and energetic QMEAN scores are present (**eFigure 2**, panels B and H), the models fall within the quality range observed in high-resolution experimental structures (**eFigure 2**, panels D and J).

**eTable 1.** Primers used for the amplification of the *MYH9* gene.

| Primer | Sequence (5’-3’) |
| --- | --- |
| MYH9-4 Reverse | CCTCAAGAATGAGAACAGACTGG |
| MYH9-9 Reverse | GGAATCATTTTCCCATACACTGAAG |
| MYH9-12 Forward | GGGCATAGGGTATGAGGGTTT |
| MYH9-12 Reverse | CCACACCCAACCAAAGTCTTCA |
| MYH9-28 Forward | TGGATCTAGGGTCCAGTGATGA |
| MYH9-28 Reverse | GCCAGTTTGAGAAGAGAGAGAGAC |
| MYH9-29 Forward | TTCTCAAACTGGCTCCTCAGAC |
| MYH9-29 Reverse | GGAGCTGGTCCTGCTGATTTA |
| MYH9-32 Forward | ATGCACGGGACTGTGTGTATT |
| MYH9-32 Reverse | TTCCAGCTGCGTCTTCATCTC |
| MYH9-33 Forward | ACGGAGATGGAGGACCTTATGA |
| MYH9-33 Reverse | CAATCCAGGTGGAAGGAGAGAAC |
| MYH9-36 Forward | CTAGAGGGTTTCTGGAGGAAGG |
| MYH9-36 Reverse | CGTTGATCAGCTCCGTGTTG |
| MYH9-37 Forward | CCCTGGCGTTAGAGGAGAAG |
| MYH9-37 Reverse | CTTCTGAACACCCAACACAGAAG |
| MYH9-38 Forward | TGGCTTCTGTGTTGGGTGTT |
| MYH9-38 Reverse | CCAACCTGTGGAAGGGATGAG |
| MYH9-39 Forward | TAAGAAGCCGGGTACACATAGG |
| MYH9-39 Reverse | CTGCTTCAGGCGGGTAGAT |

**eTable 2.** Intronic MYH9 Variants Identified in the Proband.

| **Variant** | **Intron** | **ClinVar** | **SNP** | **Proband** |
| --- | --- | --- | --- | --- |
| NM_002473.4:c333+59_333+62delinsAGC | 2 | n/a | rs372137829 | heterozygous |
| NM_002473.6(MYH9):c.1012+56C>T | 9 | 1289525 | rs3752463 | heterozygous |
| NM_002473.6(MYH9):c.1012+63G>A | 9 | n/a | n/a | heterozygous |
| NM_002473.6(MYH9):c.1554+7A>G | 13 | 44550 | rs3752462 | heterozygous |
| NM_002473.6(MYH9):c.1555-71T>C | 13 | 1243503 | rs2157257 | homozygous |
| NM_002473.6(MYH9):c.1728+9G>A | 14 | n/a | n/a | homozygous |
| NM_002473.6(MYH9):3837+25C>T | 28 | 258745 | rs4821478 | heterozygous |
| NM_002473.6(MYH9):c.5061+57C>T | 35 | 1236568 | rs13053731 | heterozygous |

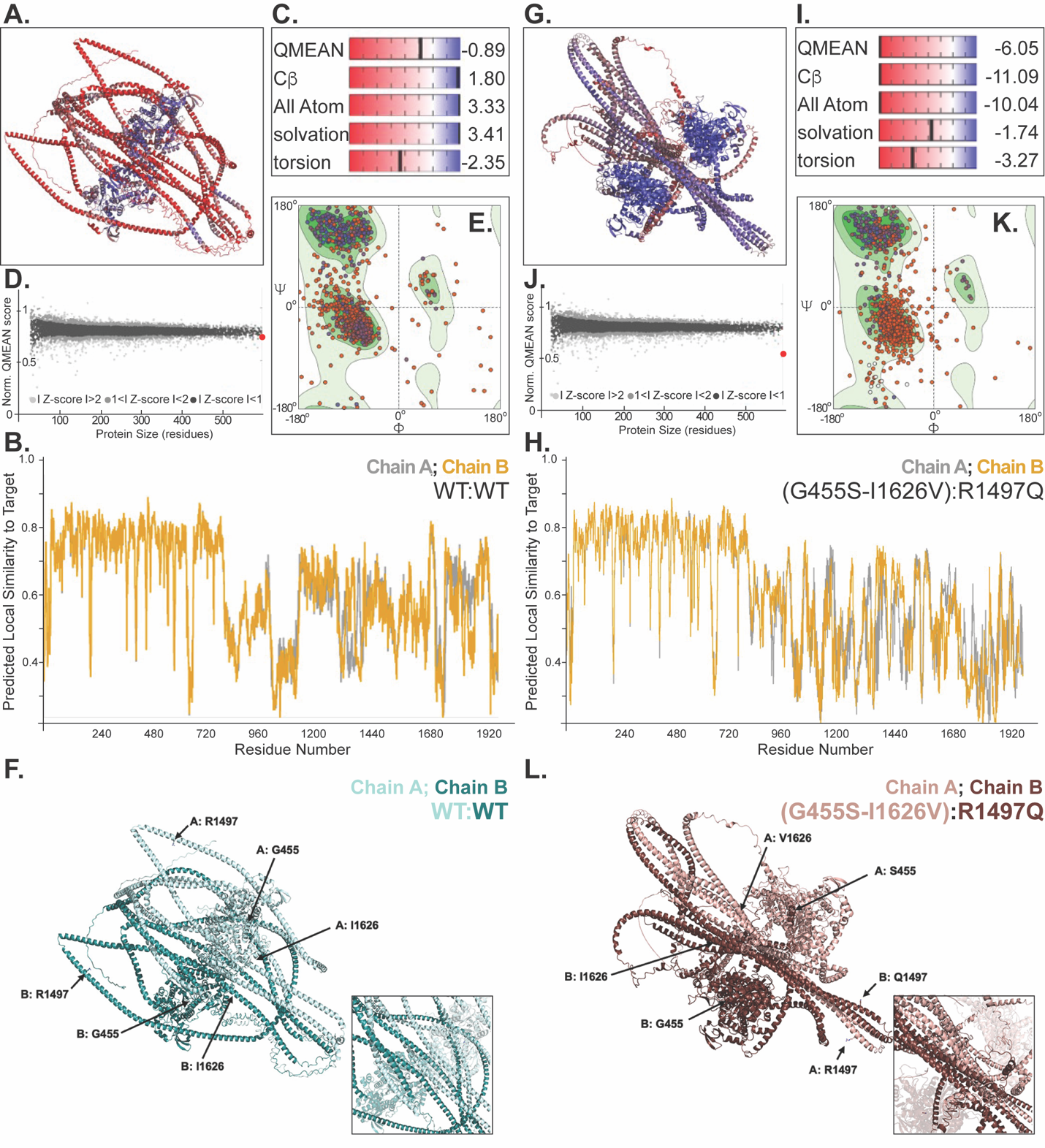

**eFigure 1. AlphaFold-Multimer *de novo* model validation and quality assessment of wild-type and mutant Myosin-9 dimer models.** Structures of the wild-type (**A**) and mutant (**G**) Gly455S-Ile1626V:Arg1497Gln [also (G455S-I1626V):R1497Q] Myosin-9 dimers, colored based on the local QMEANDisCo scores. (**B**, **H**) All Atom solvation torsion scores are depicted, contributing to QMEAN-based structural validation by evaluating solvent-accessible surface quality, atomic packing, and torsion angle geometry of the wild-type and mutant models, respectively. (**C**, **I**) Ramachandran plots illustrate the distribution of backbone dihedral angles (Φ vs Ψ) for individual residues, providing an assessment of sterically allowed conformations and local protein geometry quality. In plots E and K, 92.70% and 88.50% of residues, respectively, fall within favored regions (green), indicating high-quality structural modeling. (**D**, **J**) QMEANDisCo score provide a global and local assessment of protein model quality, with values of 0.6 (**D**) and 0.48 (**J**) for the homodimer models, respectively (indicated with a red dot). These scores are plotted against the number of residues in the homodimer, reflecting the predicted structure’s overall reliability based on protein size. (**E**, **K**) Structures of wild-type and the mutant myiosin-9 are shown, with each residue colored according to local QMEANDisCo scores in Chain A (gray) and Chain B (gold), allowing comparison of predicted structural confidence across variants. (**F**, **L**) Cartoon representation of the wild-type and mutant dimer structures highlighting the position of critical residues. Insets show central tails coils for both predicted structures.

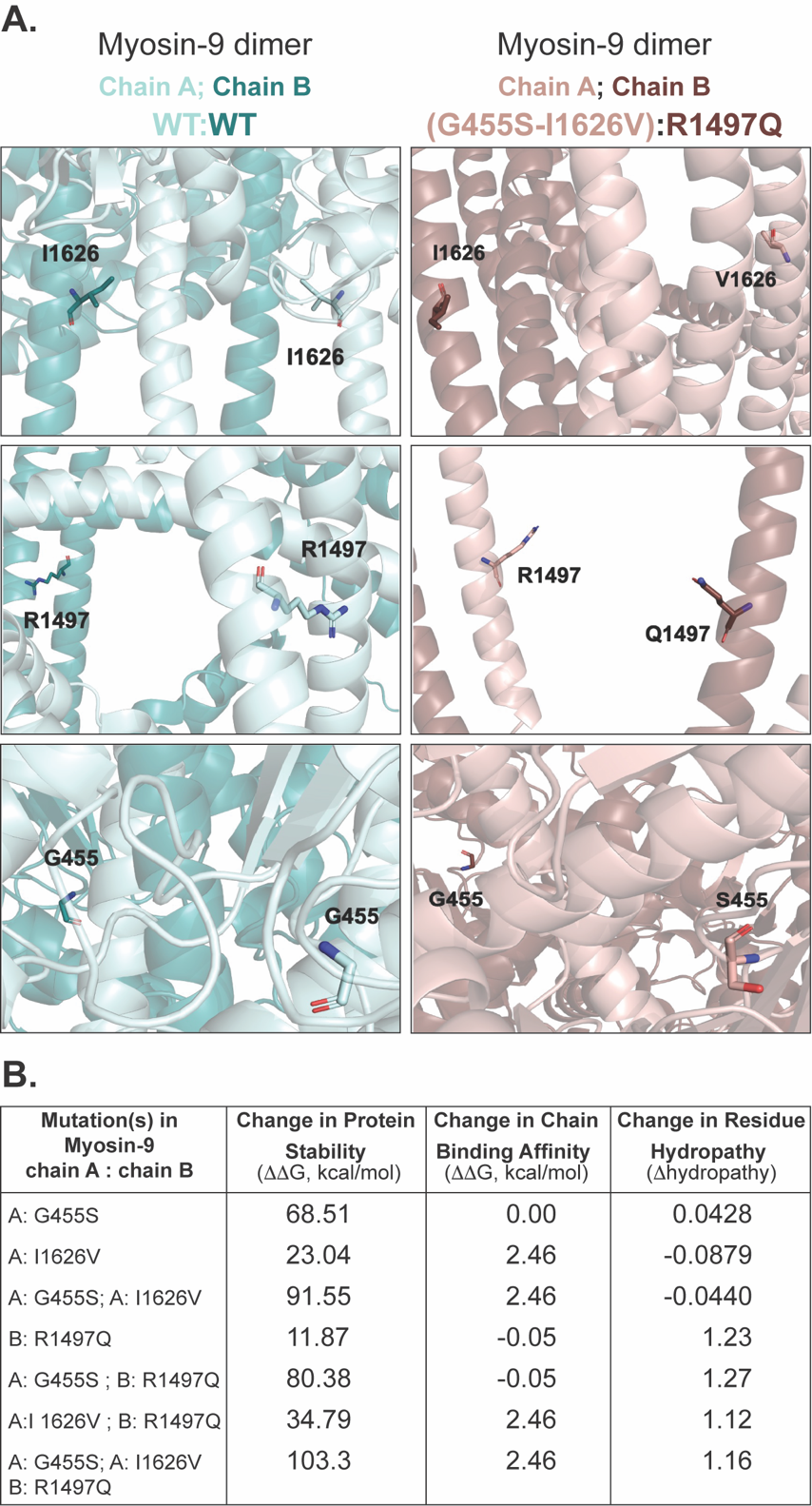

**eFigure 2.** **In silico prediction of structural and thermodynamic impacts of *MYH9* variants on Myosin-9 dimer stability and binding affinity.** AlphaFold-Multimer was used to generate the predicted Myosin-9 dimer structures (Chain A and Chain B), including the wild-type (WT) and mutated form. (**A**) Myosin-9 Gly455S-Ile1626V:Arg1497Gln [also (G455S-I1626V):R1497Q] mutations. Wild-type (teal) and mutant (brown) structures showing the interface of interactions for Gly455 (G455, top panels), Ile1626 (I1626, middle panels), and Arg1497 (R1497, lower panels). Residues are shown as sticks and colored based on atom type. (**B**) The table quantifies the impact of each mutation or combination of mutations on key protein characteristics: Mutation(s) in Myosin-9: Lists the specific amino acid changes (e.g., G455S, R1497Q, I1626V) and indicates which chain (A or B) of the Myosin-9 dimer the mutation is located on. Change in Protein Stability (ΔΔG, kcal/mol): Represents the predicted change in the overall thermodynamic stability of the protein due to the mutation (kcal/mol). Positive values typically indicate destabilization, while negative values suggest stabilization; Change in Chain Binding Affinity (ΔΔG, kcal/mol): Reflects the predicted change in the binding strength between the two Myosin-9 dimer chains (A and B) as a result of the mutation; Change in Residue Hydropathy (Δhydropathy): Indicates the predicted change in the hydropathy (hydrophobicity or hydrophilicity) of the mutated residue, with specific values provided. The table includes analyses for single mutations (e.g., A: I1626V, A: G455S, B: R1497Q) and combinations, such as the G455S and I1626V mutations on Chain A, and their occurrence with R1497Q on Chain B, reflecting complex allelic configurations
